# Recovery of intrinsic cognitive weakness in successive processing after bypass surgery for pediatric moyamoya disease

**DOI:** 10.1101/2025.02.03.25321628

**Authors:** Hideo Chihara, Takeshi Funaki, Yusuke Kusano, Yu Hidaka, Yohei Mineharu, Masakazu Okawa, Tomoki Sasagasako, Masahiro Sawada, Takayuki Kikuchi, Kanade Tanaka, Noyuri Nishida, Ami Tabata, Keita Ueda, Tsukasa Ueno, Yoshiki Arakawa

**Author notes:** **Corresponding Author:** Takeshi Funaki, MD, PhD, Department of Neurosurgery, Kyoto University Graduate School of Medicine, Kyoto, Japan 54 Shogoin Kawahara-cho, Sakyo-ku, Kyoto 606-8507, Japan.

## Abstract

**Background:** Successive processing, a form of working memory function detected with the Das Naglieri Cognitive Assessment System (CAS), is selectively impaired in pediatric moyamoya disease (MMD). We aimed to test whether successive processing in children with MMD was improved after bypass surgery under the control of confounding.

**Methods:** The present retrospective cohort study included children with MMD who underwent direct or combined bypass surgery. Neuropsychological tests including the CAS were administered at two timepoints, before and after surgery, approximately one year apart. The least squares (LS) mean standard score and LS mean difference between timepoints were calculated using a mixed model for repeated measures, which included five clinical factors along with the timepoint. Models including an interaction term were also generated to assess the effect of each clinical factor. Cognitive intra-individual variability across four domains of the CAS was assessed with an analysis of variance at each timepoint.

**Results:** Of 60 patients who underwent surgery, 42 fulfilled the inclusion criteria. The median duration between assessments was 15 months. The LS mean standard scores of successive processing increased after surgery (LS mean, 95.8 versus 100.2; LS mean difference, 4.4 [95% CI, 1.5–7.3]; *P* = 0.004). The increase was more pronounced in those with younger age at onset, shorter delay before surgery, pre-existing infarct, posterior cerebral artery involvement, and severer ischemic stage before surgery. Intra-individual variability, shown as the lowest score of successive processing at baseline, resolved after surgery (*F* = 3.56, *P* = 0.016 versus *F* = 1.21, *P* = 0.31). Successive processing was the domain most likely to be improved after surgery.

**Conclusions:** The present results suggest that successive processing is improved after bypass surgery. Larger and longer follow-up studies are required to confirm the influencing factors and long-term effect.

## Introduction

Cognitive impairment is a serious issue in children with moyamoya disease (MMD), which involves the terminal portion of the internal carotid artery (ICA) and causes cerebral ischemia. The impairment is often attributable to infarcts^1–3^; however, it can occur even with minimal radiological infarct. ^4,5^ Working memory is the most likely to be impaired among cognitive domains^6–10^ and is considered to be controlled by the prefrontal cortex. ^5,10,11^ In adult MMD, a positive effect of bypass surgery on cognitive function has been reported. ^12–14^ In pediatric MMD, however, it remains controversial whether surgery improves cognitive function. ^7,15–17^ While conservatively treated children with MMD manifested extremely poor intellectual outcomes, ^18^ pioneering observational studies reported little intellectual improvement after surgery. ^16,17^ This controversy is partly attributable to a limited number of neuropsychological tests applicable to children.

The Das Naglieri Cognitive Assessment System (CAS), a unique four-domain neuropsychological test standardized for children, can successfully characterize a distinct cognitive profile of pediatric MMD^9^; it is characterized as a selective weakness in “successive processing,” the domain reflecting verbal working memory function. ^9^ The CAS implements a calculation of “intra-individual differences,” which reflects cognitive variability, or strengths and weaknesses, across domains within an individual. ^9,19,20^ Our previous results revealed significant intra-individual differences in the domains of the CAS in children with MMD, and these differences cause underestimated difficulties in daily living. ^9^ Our more recent results suggest a correlation between low successive processing scores and reduced prefrontal blood flow (Kusano et al., in press). ^21^ However, no study has focused on the postoperative change in the CAS, especially in successive processing, as of this writing. Confounders have also rarely been adjusted in the comparison between pre- and postoperative assessments. We hypothesized that successive processing was improved after bypass surgery even with adjustment by clinical factors. The objective of our study was to compare pre- and postoperative standard scores detected with the CAS, especially focusing on the change in scores of successive processing. Our study might contribute to better understanding of the role of, and indications for, bypass surgery.

## Methods

### Data Availability Statement

Data supporting the findings of this study are available from the corresponding author upon reasonable request.

### Patients and Setting

The present study was approved by the Kyoto University Graduate School and Faculty of Medicine, Ethics Committee (R1417-1). All patients and families gave either opt-out or written informed assent/consent in accordance with the ethical guidelines for medical and health research involving human subjects in Japan. The Strengthening the Reporting of Observational Studies in Epidemiology (STROBE) statement was followed.

The present retrospective cohort study included children who were diagnosed as MMD according to the guideline^22,23^ and were admitted to Kyoto University Hospital between June 2016 and December 2023. Consecutive sampling was performed. Patients were eligible if they were aged 5 to 17 years and underwent neuropsychological testing including the CAS before surgery, underwent bypass surgery by a single surgeon (T.F.), and completed postoperative neuropsychological testing including the CAS after surgery. Patients were excluded if they did not undergo the CAS before surgery or had special developmental backgrounds judged by the psychiatrist. ^9^ Patients in whom the interval between pre- and postoperative assessments of the CAS exceeded as long as 5 years were also excluded because factors other than surgery might affect the results.

### Schedule of Neuropsychological Assessments

The schedule of neuropsychological assessments and surgical treatment is shown in Figure 1. The CAS and the Wechsler Intelligence Scale for Children Fourth Edition (WISC-IV) were routinely administered before and after surgery at intervals of 1 year. According to the manual, the interval was set to at least 6 months to minimize practice effects by repeating tests. ^20^ Administration of the tests in the acute phase of stroke was avoided. Assessments at baseline were performed during the first admission, and postoperative assessments were performed during the follow-up admission or at the outpatient clinic. Alternate forms were not used. The same occupational therapist performed both pre- and postoperative tests for each patient.

**Figure 1.**
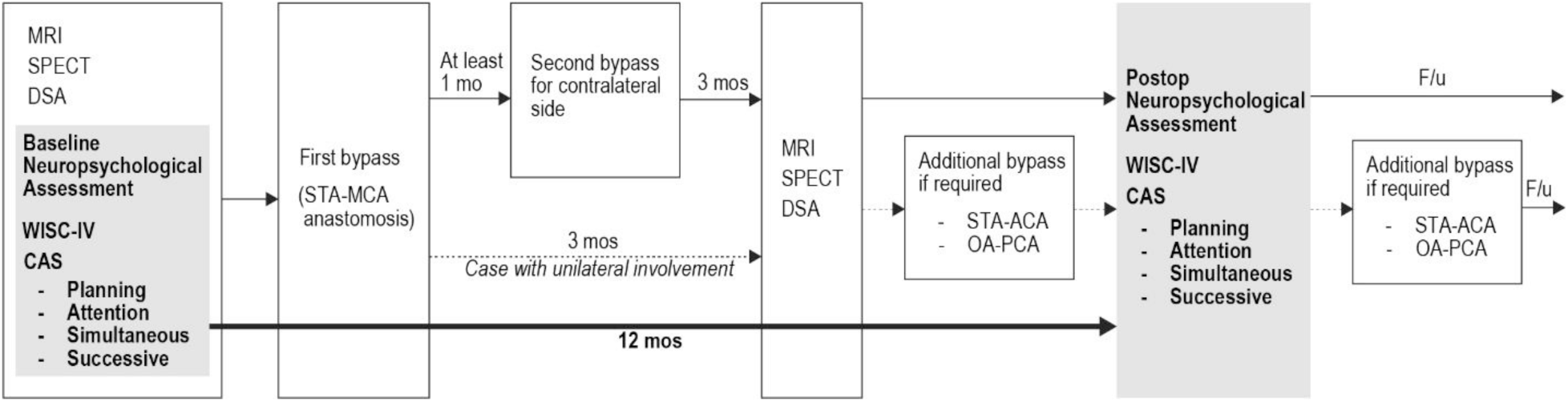
Flow chart showing the schedule of neuropsychological assessment, imaging, and surgical treatment. ACA, anterior cerebral artery; CAS, Das Naglieri Cognitive Assessment System; DSA, digital subtraction angiography; MCA, middle cerebral artery; MRI, magnetic resonance angiography; OA, occipital artery; PCA, posterior cerebral artery; STA, superficial temporal artery; SPECT, single-photon emission tomography; WISC, Wechsler Intelligence Scale for Children.

The details of the CAS have been described elsewhere. ^9^ In brief, the CAS is applicable for children aged 5 to 17 years and comprises full scale and four domains: planning, attention, simultaneous, and successive processing. The task of successive processing includes *word recall*, *sentence repetition*, *speech rate*, and *sentence question*, all of which require verbal working memory. ^9^ The standard scores of each domain are defined as a mean of 100 with a standard deviation of 15. The manual of the CAS defines the calculation of “intra-individual differences” as mentioned previously; they are calculated by subtracting the mean of the four standard scores from each standard score. ^19,20^

### Surgical Procedure

Bypass surgery was indicated for children with ischemic or hemorrhagic symptoms according to the guideline. ^24^ We have adopted conventional superficial temporal artery (STA)-middle cerebral artery (MCA) anastomosis with or without encephalomyosynangiosis as a first-line treatment. ^1,25^ For patients with bilateral ICA involvement, staged bypass surgeries for the bilateral MCA territories were performed at intervals of at least 1 month. Patients underwent follow-up imaging examinations including conventional angiography approximately 3 months after surgery and then MRI annually. During follow-up, additional revascularization of the anterior cerebral artery (ACA) or posterior cerebral artery (PCA) territory was considered if indicated. The indication criteria and procedures of revascularization of the ACA and PCA territories have been described in detail elsewhere. ^26^ In brief, additional revascularization of the ACA territory was considered if patients exhibited both remaining transient ischemic attacks (TIA) predominantly in the lower extremities and reduced CBF in the corresponding ACA territories. STA-ACA direct anastomosis combined with encephalo-pericranio-synangiosis was performed as the revascularization procedure. Similarly, additional revascularization of the PCA territory was considered if patients exhibited both transient visual symptoms and reduced CBF in the corresponding PCA territories. Occipital artery (OA)-PCA direct anastomosis combined with encephalo-pericranio-synangiosis was performed as the revascularization procedure.

### Outcome

The primary outcome variable was the standard score of successive processing, regarded as a continuous variable. The standard scores of the other domains (full scale, planning, attention, and simultaneous processing) were also evaluated.

According to the manual of the CAS, the change of the standard scores was also classified into one of the three categories: “markedly improved,” “unchanged,” and “markedly deteriorated.” The manual describes the predicted score range at the second session by each score at the first session, which is approximately ±11 points. The change was defined as “markedly improved” and “markedly deteriorated” when the second score increased and decreased beyond the predicted range, respectively. The change was defined as “unchanged” when the second score was within the predicted range.

### Clinical Factors Affecting Postoperative Effect

Clinical factors affecting outcome, in addition to bypass surgery, included the following variables: age at onset, sex, initial manifestations, radiological evidence of infarct on MRI, Suzuki’s angiographic stage, ^27^ preoperative hemodynamic stage in single-photon emission tomography (SPECT) graded as either stage 2 or non-stage 2, ^28^ laterality of disease involvement, presence or absence of PCA involvement, ^1^ and additional revascularization of the ACA or PCA territory. According to the previous study, ^1^ age at onset, delay before surgery, pre-existing radiological evidence of infarct on MRI, and PCA involvement were selected as potential confounders and incorporated into the statistical model. Preoperative SPECT stage was also incorporated into the model according to our recent results suggesting an association between successive processing and SPECT stage (Kusano et al., in press). ^21^

### Statistical Analysis

The sample size was determined by the number of cases treated during the study period. Summary statistics were constructed using frequencies and proportions for categorical data. For continuous variables, the mean and standard deviation (SD) were obtained when a distribution of normality was observed, and the median and interquartile range (IQR) when a distribution of normality was not observed. A mixed model for repeated measures (MMRM) was used for outcomes collected at two timepoints. We incorporated into the model the timepoint (baseline, postoperation), age at onset (< 6, ≥ 6 years old), delay before surgery (≥ 3, < 3 years), pre-existing radiological evidence of infarct (yes, no), PCA involvement (yes in either hemisphere, no), and preoperative SPECT stage (stage 2 in either hemisphere, non-stage 2) as fixed effects. The least squares (LS) mean at each timepoint and LS mean difference between timepoints were calculated with the corresponding two-sided 95% confidence intervals (CIs). In addition, to evaluate the effect of clinical factors that might influence postoperative change, a model incorporating an interaction term between timepoint and each clinical factor was also analyzed for each clinical factor, along with timepoint and all five clinical factors. The LS mean difference between timepoints for each clinical factor was then calculated. A one-factor repeated measures analysis of variance (ANOVA) was used to compare the standard scores across the four measures of the CAS within subject at each timepoint because the standard scores from an individual are related. ^9^ Regarding the comparison of the intra-individual differences across the four measures of the CAS, a one-way ANOVA was used. All statistical analyses were performed with JMP Pro software (version 17, SAS Institute Inc.).

## Results

### Patients

The flow diagram for patient inclusion is summarized in Figure 2. A total of 70 patients aged between 5 and 17 years underwent baseline neuropsychological testing including the CAS. Of these, 60 patients underwent bypass surgery by a single surgeon. Five patients were excluded for having a special developmental background judged by the psychiatrist, and 13 patients were excluded because they did not complete postoperative psychological testing for various reasons as listed in Figure 2. Consequently, the remaining 42 patients were analyzed.

**Figure 2.**
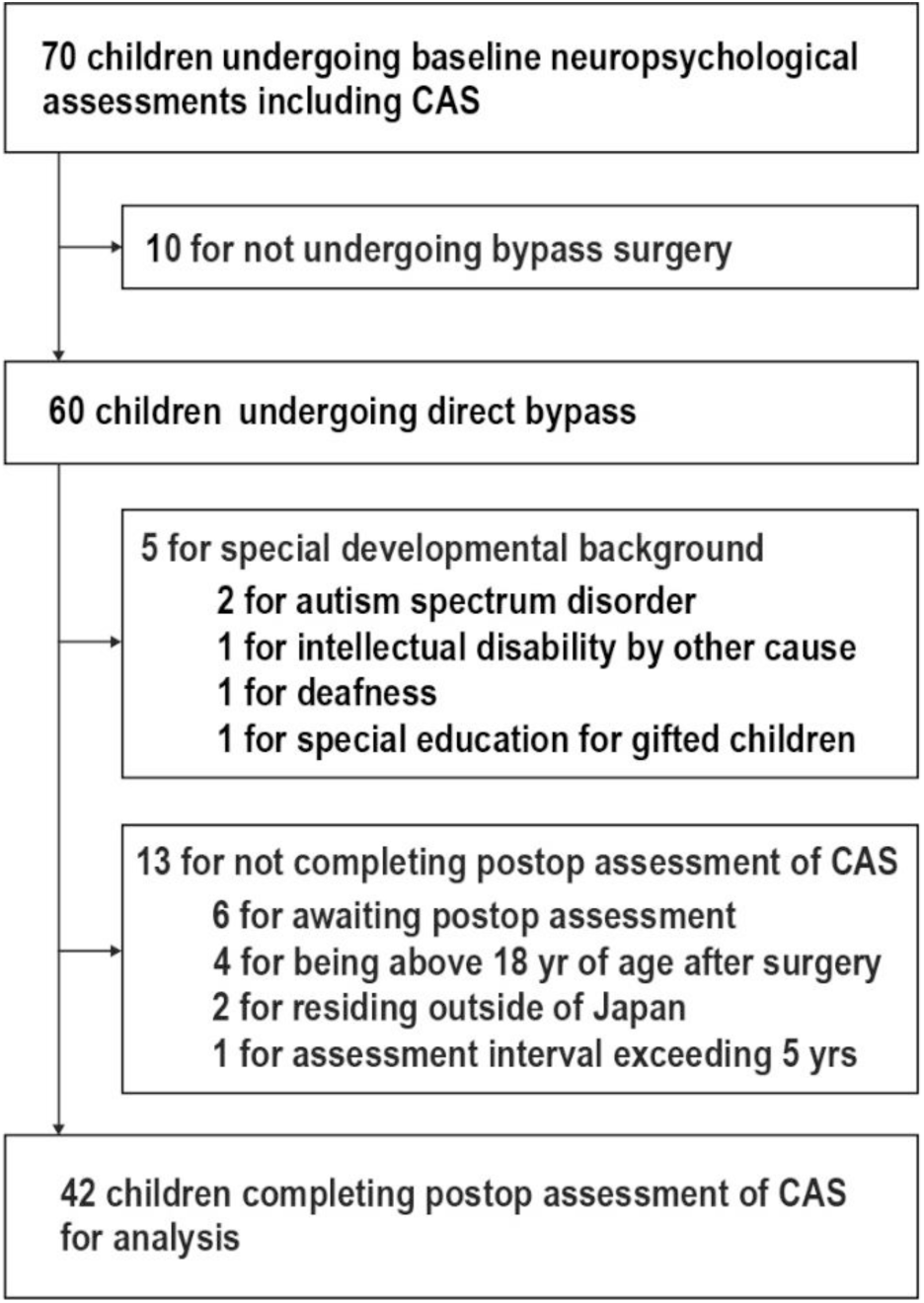
Study profile. CAS, Das Naglieri Cognitive Assessment System.

The clinical factors at baseline are summarized in Table 1. The median time interval between the baseline and postoperative assessment of the CAS was 15 months (IQR, 12.75– 20.25).

**Table 1.** Baseline Clinical Factors.

| **Variable** |  |
| --- | --- |
| No. of patients | 42 |
| Median age at onset (IQR) | 7 (6–10) |
| < 6 yrs old, n (%) | 19 (45.2) |
| ≥ 6 yrs old, n (%) | 23 (54.7) |
| Female, n (%) | 25 (59.5) |
| Initial manifestation, n (%) |  |
| TIA | 35 (83.3) |
| Others |  |
| Ischemic stroke | 1 (2.4) |
| Hemorrhagic stroke | 2 (4.7) |
| Involuntary movement | 1 (2.4) |
| Headache | 3 (7.1) |
| Radiological evidence of infarct (%) | 20 (47.7) |
| Median Suzuki stage (IQR) |  |
| Higher stage | 3 (2.75–4) |
| Right hemisphere | 3 (2–4) |
| Left hemisphere | 2 (1–3) |
| SPECT stage 2, n (%) |  |
| Either hemisphere | 23 (54.8) |
| Right hemisphere | 17 (40.5) |
| Left hemisphere | 16 (38.1) |
| Bilateral involvement (%) | 30 (71.4) |
| Right unilateral | 8 (19.0) |
| Left unilateral | 5 (11.9) |
| Presence of PCA involvement (%) | 8 (19.0) |
| Additional revascularization |  |
| ACA territory | 6 (14.3) |
| PCA territory | 1 (2.4) |
| Median interval btw baseline and postop CAS, month, (IQR) | 15 (12.75–20.25) |

ACA, anterior cerebral artery; CAS, Das Naglieri Cognitive Assessment System; IQR, interquartile range;
PCA, posterior cerebral artery; SPECT, single-photon emission tomography.

### Postoperative Change in Outcome

The baseline and postoperative standard scores in each domain of the CAS are summarized in Table 2. The LS mean standard scores of the full scale increased after surgery (LS mean, 99.6 versus 102.2; LS mean difference, 2.5 [95% CI, 0.6–4.5]; *P* = 0.011). The LS mean standard scores of successive processing also increased after surgery (LS mean, 95.8 versus 100.2; LS mean difference, 4.4 [95% CI, 1.5–7.3]; *P* = 0.004). No difference between baseline and postoperative LS mean standard scores was observed in the remaining domains: planning, attention, and simultaneous processing.

**Table 2.** Baseline and Postoperative Least Squares (LS) Mean Standard Scores in Each Domain of the Das Naglieri Cognitive Assessment System (CAS)

| Domains | Baseline<br>LS mean* | Postop<br>LS mean* | Difference (Postop – Baseline) |  |  |
| --- | --- | --- | --- | --- | --- |
|  |  |  | LS mean* | 95% CI | P |
| **Full scale** | 99.6 | 102.2 | 2.5 | 0.6, 4.5 | 0.011 |
| **Planning** | 99.6 | 97.8 | –1.8 | –5.5, 1.9 | 0.33 |
| **Attention** | 103.2 | 105.1 | 1.8 | –2.4, 6.1 | 0.39 |
| **Simultaneous** | 100.2 | 102.6 | 2.4 | –0.7, 5.5 | 0.13 |
| **Successive** | 95.8 | 100.2 | 4.4 | 1.5, 7.3 | 0.004 |

CI, confidence interval; LS, least squares
\* MMRM (mixed model for repeated measures). Model including timepoint (baseline, postop), age at onset (< 6, ≥ 6 years old), delay before surgery (≥ 3, < 3 years), radiological evidence of infarct (yes, no), PCA involvement (yes, no), and SPECT stage (stage 2, non-stage 2) as fixed effects.

### Effect of Clinical Factors

The effect of each clinical factor on the difference between baseline and postoperative successive processing scores is summarized in Table 3. Postoperative scores were larger than baseline when the age at onset was less than 6 years (LS mean difference, 7.7 [95% CI, 3.6–11.8]; *P* < 0.001), delay before surgery was less than 3 years (LS mean difference, 4.7 [95% CI, 1.5–7.9]; *P* = 0.005), preoperative infarction was present (LS mean difference, 6.1 [95% CI, 1.9–10.3]; *P* = 0.006), and either hemisphere was SPECT Stage 2 (LS mean difference, 5.7 [95% CI, 1.8–9.7]; *P* = 0.005). The postoperative scores increased for both the presence (LS mean difference, 7.0 [95% CI, 0.3–13.7]; *P* = 0.041) and absence (LS mean difference, 3.8 [95% CI, 0.5–7.0]; *P* = 0.023) of PCA involvement, but the change in scores was larger for the presence of PCA involvement.

**Table 3.** Effect of Each Confounder on Difference between Baseline and Postoperative Successive Processing Scores.

| Variable used for interaction |  | Difference (Postop – Baseline) of successive processing |  |  |
| --- | --- | --- | --- | --- |
|  |  | LS mean* | 95% CI | P |
| **Age at onset** | < 6 years | 7.7 | 3.6, 11.8 | < 0.001 |
|  | ≥ 6 years | 1.6 | –2.1, 5.3 | 0.39 |
| **Delay before surgery** | ≥ 3 years | 2.7 | –4.5, 9.9 | 0.45 |
|  | < 3 years | 4.7 | 1.5, 7.9 | 0.005 |
| **Preexisting infarct** | Yes | 6.1 | 1.9, 10.3 | 0.006 |
|  | No | 2.8 | –1.2, 6.8 | 0.16 |
| **PCA involvement** | Yes<sup>†</sup> | 7.0 | 0.3, 13.7 | 0.041 |
|  | No | 3.8 | 0.5, 7.0 | 0.023 |
| **SPECT stage** | Stage 2<sup>†</sup> | 5.7 | 1.8, 9.7 | 0.005 |
|  | Stage non-2 | 2.7 | –1.6, 7.0 | 0.20 |

CI, confidence interval; LS, least squares; PCA, posterior cerebral artery; SPECT, single-photon emission tomography
\* LS means for the interaction term using repeated measures mixed effects model (MMRM) with fixed effects for time (pre- and postoperative), age of onset, delay before surgery, infarct, PCA, SPECT and variable used for the interaction and time (pre- and postoperative).
† In either hemisphere.

### Change in Cognitive Profile

We subsequently evaluated statistical variances of the standard scores and intra-individual differences across the four domains of the CAS by each timepoint (Table 4 and Fig. 3) because cognitive intra-individual variability can cause potential difficulties in daily living. At baseline, the standard scores differed across the four domains of the CAS (*F* = 3.56, *P* = 0.016 in one-factor repeated measures ANOVA), and that of successive processing was the lowest (98.7 ± 14.8); however, these differences resolved after surgery (*F* = 1.21, *P* = 0.31). Similarly, the intra-individual differences differed across the four domains at baseline (*F* = 4.74, *P* = 0.003 in one-way ANOVA), and that of successive processing was the lowest (−4.8 ± 11.7); however, these differences resolved after surgery (*F* = 1.61, *P* = 0.19).

**Figure 3.**
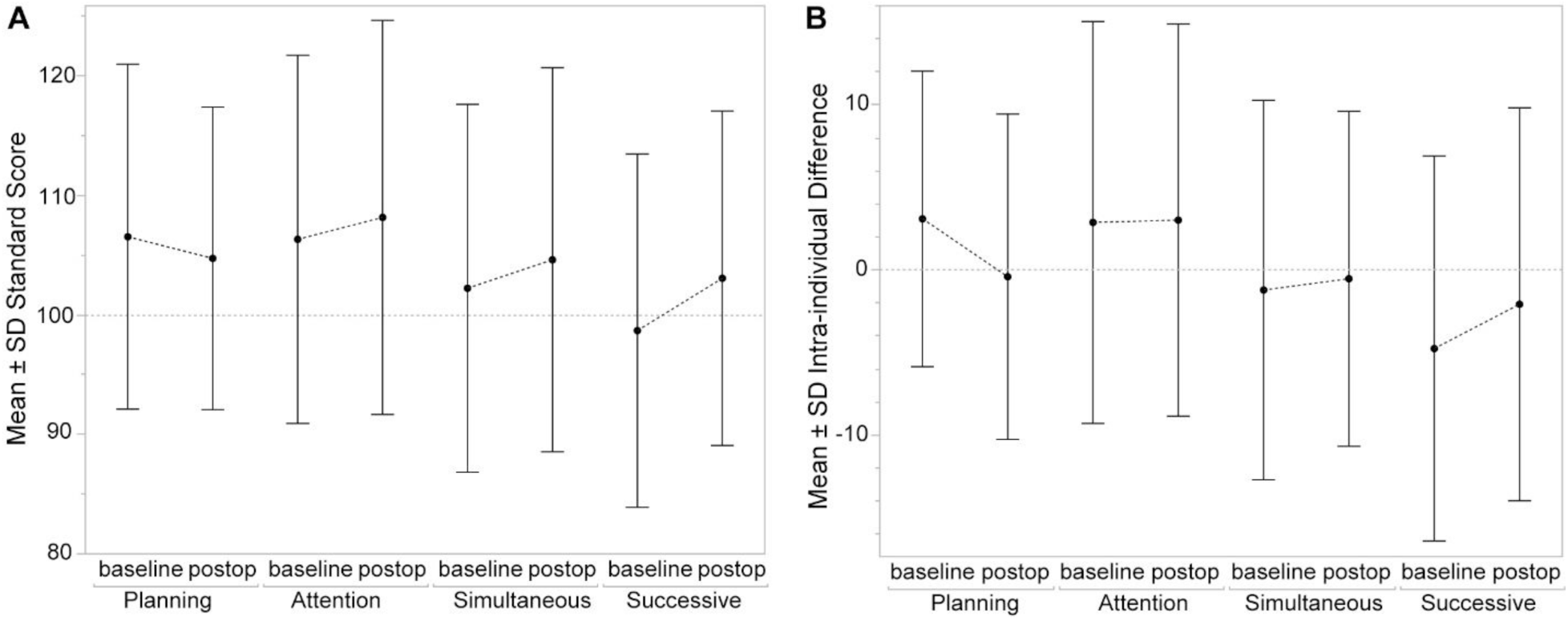
Graph showing the change in each domain of the Das Naglieri Cognitive Assessment System (CAS). **A**: Standard score. **B**: Intra-individual difference. Error bar indicates standard deviation.

**Table 4.** Standard Score and Intra-individual Difference (IID) of Each Domain of the Das Naglieri Cognitive Assessment System (CAS)

| Time-point | Planning |  | Attention |  | Simultaneous |  | Successive |  | *F* | *P*<sup>*</sup> |
| --- | --- | --- | --- | --- | --- | --- | --- | --- | --- | --- |
|  | Mean | SD | Mean | SD | Mean | SD | Mean | SD |  |  |
| **Baseline** | 106.5 | 14.4 | 106.3 | 15.4 | 102.2 | 15.4 | 98.7 | 14.8 | 3.56 | 0.016 |
| **Postop** | 104.7 | 12.7 | 108.2 | 16.5 | 104.6 | 16.1 | 103.1 | 14.0 | 1.21 | 0.31 |
|  | IID Mean | SD | IID Mean | SD | IID Mean | SD | IID Mean | SD | *F* | *P*<sup>†</sup> |
| **Baseline** | 3.1 | 9.0 | 2.9 | 12.2 | -1.2 | 11.5 | -4.8 | 11.7 | 4.74 | 0.003 |
| **Postop** | -0.4 | 9.9 | 3.0 | 11.9 | -0.5 | 10.2 | -2.1 | 11.9 | 1.61 | 0.19 |

IID, intra-individual difference; SD, standard deviation.
\* One-factor repeated measures ANOVA
† One-way ANOVA

The category of the change in each domain, defined according to the manual, is shown in Figure 4. Regarding successive processing, 9 (21.4%) of 42 patients were classified as “markedly improved,” 32 (76.2%) as “unchanged,” and 1 (2.4%) as “markedly deteriorated.” Successive processing was the most likely to be improved and the least likely to deteriorate among domains (Fig. 4).

**Figure 4.**
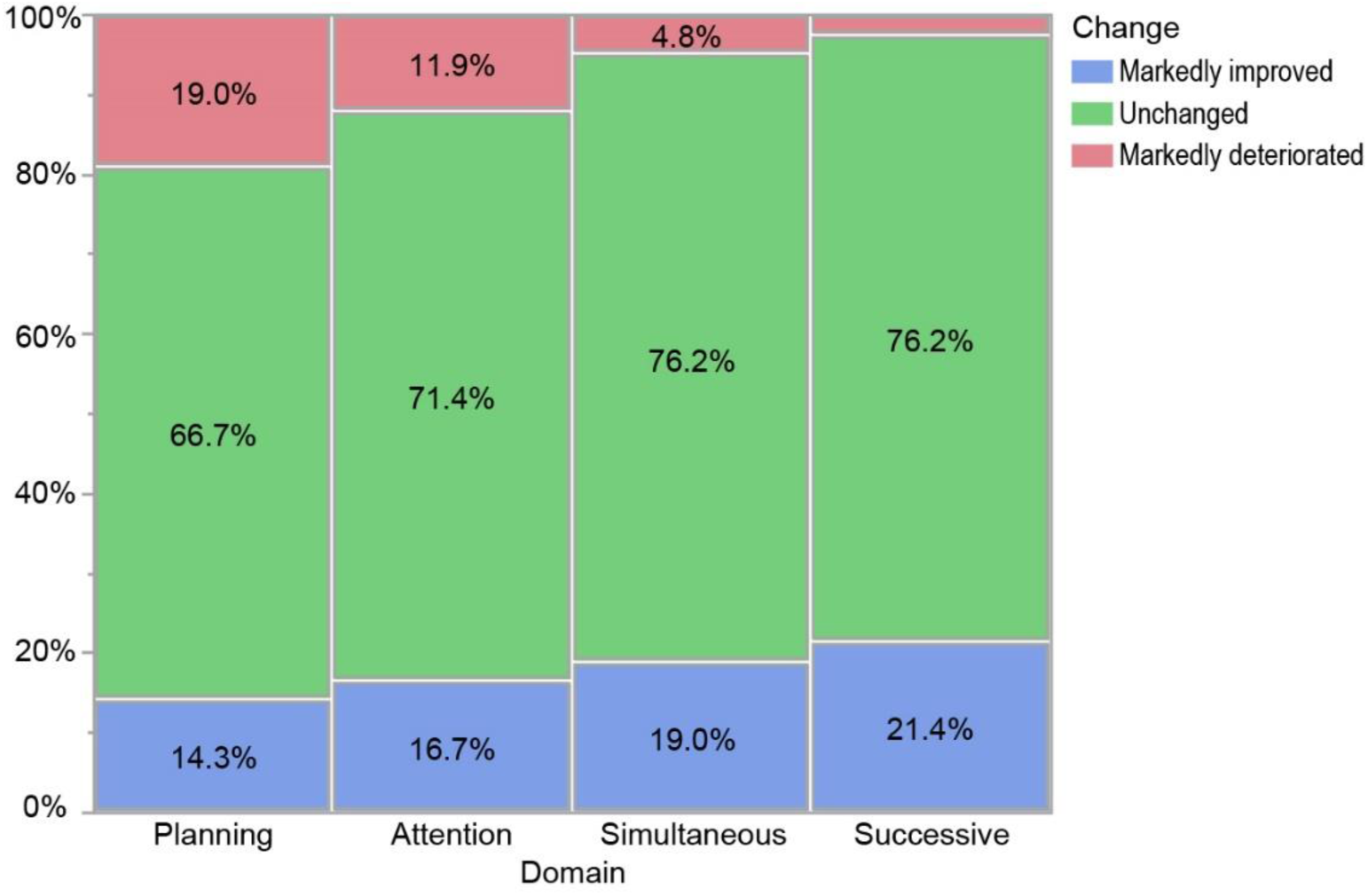
Graph showing the category of the change in each domain of the Das Naglieri Cognitive Assessment System (CAS).

## Discussion

In addition to the previous studies on postoperative improvement of neurocognitive function, our study has three key findings. First, the standard score of successive processing increased after surgery under the control of confounding. Second, the improvement in successive processing score was affected by several clinical factors including age at onset, delay before surgery, pre-existing infarct, posterior cerebral artery involvement, and ischemic stage before surgery. Third, cognitive intra-individual variability, shown as the lowest score of successive processing among domains at baseline, resolved after surgery. The present study is perhaps the first to reveal the cognitive improvement detected with the CAS after bypass surgery for pediatric MMD.

Our results are in line with those of previous studies suggesting the beneficial effect of bypass surgery on cognitive function in MMD. Kimura et al. analyzed the pre- and postoperative neuropsychological tests in 52 adult patients and found an increase of scores in verbal and performance intelligence quotients (IQs) in the Wechsler Adult Intelligence Scale-Revised (WAIS-R), memory quotient of the Wechsler Memory Scale, and Rey copy and recall tests. ^12^ Two other studies on adult patients also suggested improvements in verbal and performance IQs in WAIS in the group in which cerebral blood flow was ameliorated after bypass surgery. ^13,14^ Regarding pediatric MMD, Lee JY et al. analyzed the pre- and postoperative cognitive profiles of 65 patients using WISC-R. They found that performance IQ, especially the Coding subtest, increased after surgery. Coding tasks measure processing speed and reflect various cognitive functions including working memory. Our results add novel information to these seminal studies because successive processing, a cognitive function intrinsically impaired in pediatric MMD, ^9,21^ was improved after surgery.

On the other hand, our results are partly inconsistent with a historical work by Matsushima Y et al., in which full-scale and performance IQ in WISC were deteriorated during follow-up after indirect bypass surgery. ^16^ Digit span, which measures working memory, was the most likely to be deteriorated among subtests. ^16^ Several possible reasons for this inconsistency are considered, including differences in surgical procedures and length of follow-up period. Another possible reason is that the tasks of successive processing of the CAS might sensitively detect cognitive improvement after surgery, considering that the CAS is a sensitive method for detecting cognitive characteristics in pediatric MMD. ^9^

The improvement of successive processing shown in the present study is reasonably explained by the probable association between working memory function and cerebral blood flow especially in the prefrontal cortex. Karashima et al. revealed that the working memory index in WISC was associated with blood flow in the anterior area of the middle cerebral artery territory. ^11^ In adult MMD, neuronal loss in the medial frontal lobe, which is detected with ^123^I-iomazenil SPECT, was related to cognitive dysfunction. ^5^ Kuroda et al. showed that the type of bypass surgery which widely covered the frontal lobe was a factor associated with favorable intellectual outcomes. ^29^ Our previous study also revealed that selective intra-individual weakness in successive processing was associated with reduced blood flow in the prefrontal cortex. ^21^ All these findings suggest that the amelioration of blood flow in the prefrontal cortex results in the recovery of successive processing; however, this speculation should be confirmed by further studies.

We observed that the postoperative increase of successive processing scores was more pronounced in patients who underwent surgery within 3 years after symptom onset than it was in those with a longer delay (Table 3). Viewed from a clinical perspective, this may suggest that early surgical intervention is recommended for children with MMD in terms of cognitive recovery. This speculation is supported by the study by Imaizumi et al., in which longer duration after onset without surgery was associated with unfavorable intellectual outcomes. ^30^ Larger studies are required to identify factors affecting the improvement of successive processing.

Our study has several limitations. First, the study was a retrospective one and did not have a control group not undergoing bypass surgery. However, our conclusions might be justified considering the extremely poor intellectual outcomes in conservatively treated children found by an earlier study. ^18^ Second, the sample size of the present study was not sufficient to conduct a logistic regression analysis identifying factors associated with the improvement of successive processing as discussed previously. Third, we observed postoperative cognitive improvement at the median interval of 15 months; however, it remains unclear whether the improvement lasts for a long time after surgery. As shown in Table 4 and Figure 3, the score of successive processing remained the lowest among domains even after surgery. Long-term careful follow-up is required, and adequate educational support should be considered for those with lasting weakness of successive processing.

## Conclusions

The present results support our hypothesis that successive processing, a working memory function intrinsically impaired in pediatric MMD, is improved after bypass surgery. Larger studies are required to identify factors affecting the improvement. Long-term follow-up is also required to confirm whether the improvement lasts for a long time after surgery.

## Nonstandard Abbreviations and Acronyms

CAS: the Das Naglieri Cognitive Assessment System
MMD: moyamoya disease.

## Acknowledgments

None.

